# “Time does not heal all wounds”: Sexual victimisation is associated with depression, anxiety and PTSD in old age

**DOI:** 10.1101/2021.06.05.21258397

**Authors:** Anne Nobels, Gilbert M.D. Lemmens, Lisa Thibau, Marie Beaulieu, Christophe Vandeviver, Ines Keygnaert

## Abstract

**Background:** Sexual violence (SV) has an important impact on mental health. Childhood sexual abuse is linked to internalising disorders in later life. In older adults, SV occurs more often than previously believed. Moreover, health care workers lack the skills to address SV in later life. Studies researching the mental health impact of lifetime SV, i.e. SV during childhood, adulthood and old age, are lacking.

**Methods:** Between July 2019 and March 2020, 513 older adults living in Belgium participated in structured face-to-face-interviews. Selection occurred via a cluster random probability sampling with a random walk finding approach. Depression, anxiety and posttraumatic stress syndrome (PTSD) were measured using validated scales. Participants were asked about suicide attempts and self-harm during their lifetime and in the past 12-months. SV was measured using behaviourally specific questions based on a broad SV definition.

**Results:** Rates for depression, anxiety and PTSD were 27%, 26% and 6% respectively, 2% had attempted suicide, 1% reported self-harm in the past 12-months. Over 44% experienced lifetime SV, 8% in the past 12-months. Lifetime SV was linked to depression (p =.001), anxiety (p =.001), and PTSD in participants with a chronic illness/disability (p = .002) or no/lower education (p <.001). We found no link between lifetime SV and suicide attempts or self-harm in the past 12-months.

**Conclusions:** Lifetime SV is linked to mental health problems in late life. Tailored mental health care for older SV victims is necessary. Therefore, capacity building of professionals, and development of clinical guidelines and care procedures are important.

## 1. Introduction

Sexual violence (SV) (WHO, 2017a) is increasingly considered an important public health problem of major societal concern (World Health Assembly, 1996). In older adults, SV has been studied in the broader context of elder abuse and neglect for many years. Moreover, most studies only included hands-on SV (e.g. unwanted kissing, rape), whereas hands-off SV (sexual harassment without physical contact) was not studied. Therefore, prevalence numbers have been underestimated for a long time (Nobels et al., 2020). A recent study, which assessed SV independently from other forms of violence and used a broad definition of SV encompassing both hands-off and hands-on SV, showed that SV in older adults occurs more frequently than previously believed (Nobels et al. 2021a).

In older adults, mental health problems are common and lead to impaired functioning in daily life (WHO, 2017b). Research has shown that up to one in three older adults reports depressive symptoms (Meeks, Vahia, Lavertsky, Kulnarni, & Jeste, 2011), up to 14% suffers from an anxiety disorder (Wolitzky‐Taylor, Castriotta, Lenze, Stanley, & Craske, 2010), and 3% from PTSD (Böttche, Kuwert, & Knaevelsrud, 2012). Furthermore, older adults complete suicide proportionally more often than any other age group (Rodda, Walker, & Carter, 2011). Physical and verbal abuse against older adults have been associated with adverse mental health outcomes (Olofsson, Lindqvist, & Danielsson, 2012). When it comes to SV, studies tend to focus only on the long-term mental health outcomes of child sexual abuse (CSA), indicating that CSA victims suffer more from internalising disorders in later life (Ege, Messias, Thapa, & Krain, 2015; Rapsey, Scott, & Patterson, 2019). However, studies taking a life course perspective and researching the mental health impact of lifetime sexual victimisation, including SV that happened during childhood, adulthood, and old age, are currently lacking. Several risk factors have been associated with both elder abuse and neglect and adverse mental health outcomes in late life such as being female, increasing age, low education level, financial difficulties, suffering from a chronic illness or disability, being care dependent, low social support, and lacking resilience (Böttche et al., 2012; Luoma et al., 2011; Naughton et al., 2012; Rodda et al., 2011; Wolitzky‐Taylor et al., 2010). Both being resilient and having a high level of social support have already been shown to decrease the mental health impact of sexual victimisation in younger populations (Declercq & Palmans, 2006; Declercq, Vanheule, Markey, & Willemsen, 2007; Fedina et al., 2021; Schulz et al., 2014). Moreover, being non-heterosexual has been associated specifically with SV in older adults and has also been linked to mental health problems in older age (Nobels et al., 2021a; Yarns, Abrams, Meels, &Sewell, 2016).

Additionally, a recent study showed that sexual victimisation remains largely undetected by mental health care workers (Nobels et al., 2021b). Although the World Psychiatric Association (WPA) petitioned a routine inquiry on SV in all psychiatric assessments (Steward & Chandra, 2017), health care workers lack the appropriate communication skills to adequately address SV in later life. They are worried that talking about SV will break down the victims’ defence mechanisms and that they will feel shocked, and helpless when confronted with SV disclosure by older victims (Goldblatt, Band-Winterstein, Lev, & Harel, 2020).

In order to provide tailored care to older victims of SV, a better understanding of the mental health impact of lifetime sexual victimisation is needed. To our knowledge, this study is the first to assess the impact of lifetime sexual victimisation on mental health outcomes in a representative sample of older adults. We used a broad definition of SV and included sexual victimisation across the life course, including SV that happened during childhood, adulthood, and old age. This study was a part of the first gender- and age-sensitive SV prevalence study in Belgium, which aimed for a better understanding of the mechanisms, nature, magnitude, and impact of SV (Keygnaert et al., 2018). The objectives of this paper are three-fold: (1) to establish the prevalence of depression, anxiety, PTSD, suicide attempts and self-harm and its moderators in older adults in Belgium, (2) to research whether lifetime sexual victimisation is associated with depression, anxiety, PTSD, suicide attempts and self-harm in older adults, (3) to test the moderating effect of the previous identified moderators of elder abuse and neglect, SV and mental health problems (i.e. past 12-months SV, sociodemographic characteristics, health status, social support, and resilience), in the relation between lifetime sexual victimisation and depression, anxiety, and PTSD in older adults.

Based on our results we formulate recommendations for future research and health care practices.

## 2. Methods

### 2.1 Sample selection

Between 8^th^ July 2019 and 12^th^ March 2020, a representative sample of the Belgian older population participated in a structured face-to-face interview. Participants were selected using a cluster random probability sampling with a random walk finding approach (Nobels et al., 2021c). They had to live in Belgium, speak Dutch, French or English, be at least 70 years old, and have sufficient cognitive ability to complete the interview. Cognitive status was not formally tested but it was assessed based on the ability to maintain attention during the interview and the consistency of the participant’s answers, by means of a control question comparing the participant’s birth year and age. Structured face-to-face interviews were carried out in private at the participant’s place of residence by trained interviewers. Both older adults living in the community and living in nursing homes or assisted living facilities were included. In total, 513 interviews were included in the analysis. Participation rate was 34%. The full study protocol is available elsewhere (Nobels et al., 2021c).

The authors assert that all procedures contributing to this work comply with the ethical standards of the relevant national and institutional committees on human experimentation and with the Helsinki Declaration of 1975, as revised in 2008. The study received ethical approval from the ethical committee of Ghent University/University Hospital (B670201837542) and was conducted according to the WHO ethical and safety recommendations for SV research (WHO, 2016). As recommended by these guidelines, the study was presented as the “Belgian study on health, sexuality and well-being”. Written informed consent was obtained from all participants. After participation participants received a brochure with the contact details of several helplines.

### 2.2 Definitions and measures

This study was a part of a national SV prevalence study in the Belgian population between 16 and 100 years old (Keygnaert et al., 2018). The questionnaire consisted of several modules including sociodemographic characteristics, sexual health & relations, mental health, and sexual victimisation. The questions in these modules were identical across all age groups.

Mental health was measured using international scales which were validated in several age groups. Depression was measured using the Patient Health Questionnaire (PHQ)-9 (Kroenke & Spitzer, 2002) (Cronbach’s alpha (α) =0.737) and anxiety using the General Anxiety Disorder (GAD)-7 (Löwe et al., 2008) (α=0.827). Both scales assessed symptoms in the two weeks before the interview. PTSD was assessed with the Primary Care PTSD Screen for DSM-5 (PC-PTSD-5) (Prins, Scott, & Patterson, 2015) (α= 0.572), which questioned symptoms in the month before the interview. Resilience was estimated using the Brief Resilience Scale (BRS) (Smith et al., 2008) (α= 0.821). Additionally, participants were asked about self-harm and suicide attempts, both during their lifetime and in the past 12-months. Social support was measured by number of confidants. SV was defined according to the WHO definition which encompasses different forms of sexual harassment without physical contact, sexual abuse with physical contact but without penetration and (attempted) rape (WHO, 2017a). As a result of recent insights in the field of SV in older adults, this definition was expanded to include sexual neglect (Nobels et al., 2020), which was measured as ‘touching in care’ due to the absence of a standardised measure (See Appendix 1). We used behaviourally specific questions (BSQ) to assess lifetime and past 12-months SV experiences. The SV items were based on existing surveys (Keygnaert et al., 2015; Koss & Gidycz, 1985; Krahé et al., 2015), and adapted to the Belgian social and legal context (Depraetere et al., 2020). To calculate lifetime and past 12-months sexual victimisation, we created dichotomous variables out of all SV items. Due to a high level of multicollinearity between hands-off and hands-on SV (Variance Inflation Factor (VIF) >2.5) (Allison, 2012), they were combined into the variables lifetime and past 12-months SV. A detailed overview of the SV outcome measures can be found in Appendix 1.

### 2.3 Statistical analysis

Statistical analysis was performed using SPSS Statistics 26 and R version 3.6.3. Descriptive statistics (means, standard deviations, and percentages) were calculated for all variables. Outcome variables were described in a categorical way as this is how the scales are used in clinical practice. We conducted a bivariate analysis to compare the proportion of sexual victimisation within the different categories of depression, anxiety, and PTSD using a Chi-square-test or a Fisher’s exact test if the assumptions of the Chi-square-test were not met.

To assess the association between lifetime sexual victimisation and depression, anxiety, and PTSD we performed stepwise linear regression analyses for each outcome variable separately. In each model we used the score on the PHQ-9, the GAD-7, and the PC-PTSD-5 scale as a continuous outcome variable. In each analysis main effects were added and/or deleted stepwise based on selection criteria (p < 0.05). The multi-collinearity assumption of multivariate regression analyses was tested for the main effects with the VIF and indicated no violation. Given the low numbers for suicide-attempts and self-harm in the past 12-months we did not perform a regression analysis on these variables.

We constructed cross-product terms to further explore the moderating effects of past 12-months SV, sex at birth, age, sexual orientation, resilience, education level, perceived health status, care dependency, and social support, in the relation between lifetime sexual victimisation and depression, anxiety, or PTSD. The potential moderators were dichotomised (see Table 3). Continuous variables were dichotomised based on the mean value. Afterwards cross-product terms were created between each of the binary moderators and lifetime SV. Again, stepwise linear regression analyses were conducted. In each analysis the cross-product terms were added and/or deleted stepwise together with their main effects based on selection criteria (p < 0.10). Interactions were further explored if p < 0.10 to account for possible underpowering of the sample. In that case separate regression analyses were run to further analyse the interaction effect (e.g. separate analyses for higher versus no/lower education). In the final model only significant (p < 0.05) main effects and interaction effects were retained. P-values, coefficients and 95% confidence intervals (CI) are presented.

## 3. Results

### 3.1 Descriptive statistics

The study sample consisted of a representative sample of the Belgian population aged 70 years and older (n=513) (Nobels et al., 2021c). The mean age was 79 years (SD: 6.4yrs, range 70-99yrs), 58.3% was female, 7.4% labelled themselves as non-heterosexual, 89.8% was community-dwelling, 31.2% had completed higher education, 26.1% labelled their financial situation as difficult and 48.3% reported low social support. Regarding health status, 45.8% indicated that they suffered from an illness or disability limiting their daily activities, 46.4% was care-dependent and 26.5% reported low resilience. Over 44% of the participants experienced SV during their lifetime, 8.4% in the past 12-months. A detailed description of the different forms of sexual victimisation has been published elsewhere (Nobels et al. 2021a). More information on the characteristics of the study sample can be found in Table 1.

**Table 1.** Descriptive statistics of sociodemographic characteristics, health status, social support, resilience and sexual victimisation (n=513)

|  | Variable | n (%) |
| --- | --- | --- |
| Sex at birth | Female | 299 (58.3) |
|  | Male | 214 (41.7) |
| Age<sup>a</sup><br>(mean 79yrs) | 70-79yrs | 283 (55.2) |
|  | 80-89yrs | 201 (39.2) |
|  | 90-99yrs | 29 (5.7) |
| Sexual orientation | Heterosexual | 475 (92.6) |
|  | Non-heterosexual<sup>b</sup> | 38 (7.4) |
| Living situation | Community-dwelling | 461 (89.8) |
|  | Assisted living facility | 25 (4.9) |
|  | Nursing home | 27 (5.3) |
| Education level | No/lower education<sup>c</sup> | 353 (68.8) |
|  | Higher education | 160 (31.2) |
| Financial status<sup>d</sup> | Easy | 377 (73.5) |
|  | Difficult | 134 (26.1) |
| Social support<sup>e</sup> | Low | 246 (48.3) |
|  | High | 265 (51.7) |
| Perceived health status | No disability/chronical illness | 278 (54.2) |
|  | Disability/chronical illness | 235 (45.8) |
| Care dependency | Yes | 238 (46.4) |
|  | No | 275 (53.6) |
| Resilience<sup>f</sup> | Low | 136 (26.5) |
|  | Normal | 319 (62.2) |
|  | High | 58 (11.3) |
| Sexual victimisation | Lifetime | 227 (44.2) |
|  | Past 12-months | 43 (8.4) |

<sup>a</sup>Calculated based on the participant's year of birth.
<sup>b</sup>This group contains participants who labelled themselves as: homosexual, bisexual, pansexual, asexual or other. In this last group, several participants labelled themselves as "normal". Since it was not clear whether they had difficulties understanding the different terms defining sexual orientation or whether they indeed labelled their sexual orientation as "other", we decided to classify these participants as non-heterosexual based on their answer.
<sup>c</sup>This group contains participants who had no formal education or completed primary education, secondary education, technical or vocational education or religious school.
<sup>d</sup>Measured through the ability to make ends meet. The four answer categories 'with great difficulty', 'with some difficulty', 'fairly easily' and 'easily' were dichotomised into 'difficult' and 'easy'.
<sup>e</sup>Measured by number of confidants. The average number of confidants was 3. Fewer than 3 confidants corresponds to low social support, 3 or more confidants corresponds to high social support.
<sup>f</sup>Measured using the Brief Resilience Scale (BRS): Low (1.00-2.99); Normal (3.00-4.30); High (4.31-5.00)

Regarding mental health status (see Table 2), 19.9% reported mild, 5.1% moderate, and 2.5% severe depressive symptoms; and 17.5% mild, 4.5% moderate, and 3.5% severe anxiety symptoms during the two weeks prior to the interview. During the month prior to the interview, 5.7% suffered from symptoms of PTSD. We found a significant difference in level of depression, anxiety and PTSD between SV victims and non-victims (p < 0.05). Over 5% attempted suicide during their lifetime, 1.6% in the past 12 months, 2.1% reported self-harm during their lifetime, 1.4% in the past 12-months. For both suicide attempts and self-harm we found no significant difference between SV victims and non-victims.

**Table 2:** Descriptive statistics of depression, anxiety, PTSD, suicide attempts, self-harm and sexual victimisation

| Item | Scale | Outcome | N total | % total | % SV | Chi<sup>2</sup>-test |
| --- | --- | --- | --- | --- | --- | --- |
| Depression<br>(n = 512) | PHQ-9<br>($\alpha = .737$) | No | 371 | 72.5 | 41.2 | p = .047 |
|  |  | Mild | 102 | 19.9 | 47.1 |  |
|  |  | Moderate | 26 | 5.1 | 65.4 |  |
|  |  | Moderately severe/<br>Severe | 13 | 2.5 | 61.5 |  |
| Anxiety<br>(n = 513) | GAD-7<br>($\alpha = .827$) | No | 382 | 74.5 | 40.8 | p = .043 |
|  |  | Mild | 90 | 17.5 | 51.1 |  |
|  |  | Moderate | 23 | 4.5 | 60.9 |  |
|  |  | Severe | 18 | 3.5 | 61.1 |  |
| PTSD<br>(n = 512) | PC-PTSD-5 | Yes | 29 | 5.7 | 65.5 | p = .018 |
| Suicide attempts<br>(n=513) | NA | Lifetime | 27 | 5.3 | 51.9 | p = .414 |
|  |  | Past 12-months | 8 | 1.6 | 37.5 | p = 1.00 |
| Self-harm<br>(n=512) | NA | Lifetime | 11 | 2.1 | 36.4 | p = .762 |
|  |  | Past 12-months | 7 | 1.4 | 28.6 | p = .471 |

$\alpha$ = Cronbach's Alfa, SV= sexual victimisation, PTSD= Posttraumatic Stress Disorder
% SV: proportion of sexual victimisation within the different categories of depression, anxiety, PTSD, suicide attempts and self-harm
Patient Health Questionnaire-9 (PHQ-9): Mild (5-9), Moderate (10-14), Moderately severe – Severe ( $\geq 15$ )
General Anxiety Disorder-7 (GAD-7): Mild (5-9), Moderate (10-14), Severe ( $\geq 15$ )
Primary Care PTSD Screen for DSM-5 (PC-PTSD-5): Yes ( $\geq 3$ )

### 3.2. Association between lifetime SV and depression, anxiety and PTSD

The linear regression analysis (Table 3) showed that having experienced lifetime SV, financial difficulties, having a chronic illness or disability limiting daily activities, being care dependent, and having fewer than three confidants were associated with depression in old age. Exposure to SV in the past 12-months, sex at birth, age, sexual orientation, education level, and resilience were not associated with depression in our sample.

**Table 3.** Association of lifetime sexual victimisation with depression, anxiety and PTSD

|  | Depression<br>(PHQ-9) |  |  | Anxiety<br>(GAD-7) |  |  | PTSD<br>(PC-PTSD-5) |  |  |
| --- | --- | --- | --- | --- | --- | --- | --- | --- | --- |
|  | Coeff. | 95% CI | p-value | Coeff. | 95% CI | p-value | Coeff. | 95% CI | p-value |
| **Main effects** |  |  |  |  |  |  |  |  |  |
| Lifetime SV | 1.074 | .451, 1.697 | .001 | 1.173 | .504, 1.841 | .001 | .284 | .129, .439 | <.001 |
| Education level | -- | -- | -- | -.880 | -1.615, -.145 | .019 | .039 | -.129, .208 | .647 |
| Financial status | 1.248 | .544, 1.953 | .001 | 1.287 | .518, 2.056 | .001 | .211 | .034, .388 | .019 |
| Perceived health status | 1.988 | 1.324, 2.651 | <.001 | 1.345 | .633, 2.057 | <.001 | .225 | .069, .380 | .005 |
| Care dependency | 1.210 | .548, 1.872 | <.001 | 1.259 | .544, 1.974 | .001 | -- | -- | -- |
| Social support | .823 | .201, 1.445 | .010 | .990 | .320, 1.659 | .004 | -- | -- | -- |
| Resilience | -- | -- | -- | .751 | .075, 1.426 | .029 | -- | -- | -- |
| **Cross-product terms** |  |  |  |  |  |  |  |  |  |
| Lifetime SV * education level | -- | -- | -- | -- | -- | -- | .318 | -.017, .652 | .062 |
| Lifetime SV * perceived health status | -- | -- | -- | -- | -- | -- | .310 | .000, .620 | .050 |
| **Interaction effects** |  |  |  |  |  |  |  |  |  |
| Lifetime SV by education level | -- | -- | -- | -- | -- | -- |  |  |  |
| Association in higher education | -- | -- | -- | -- | -- | -- | .101 | -.178, .380 | .477 |
| Association in no/lower education | -- | -- | -- | -- | -- | -- | .356 | .165, .547 | <.001 |
| Lifetime SV by perceived health status | -- | -- | -- | -- | -- | -- |  |  |  |
| Association in no chronic illness/disability | -- | -- | -- | -- | -- | -- | .154 | -.026, .334 | .093 |
| Association in chronic illness/disability | -- | -- | -- | -- | -- | -- | .422 | .155, .689 | .002 |

SV= sexual victimisation, PTSD= Posttraumatic Stress Disorder, PHQ-9= Patient Health Questionnaire-9, GAD-7= General Anxiety Disorder-7, PC-PTSD-5= Primary Care PTSD Screen for DSM-5, Coeff= Coefficient, CI= Confidence Interval.
The coefficient describes the variability of the outcome variable in comparison to a reference value. For lifetime and past 12-months SV this is "no SV", for Sex at birth this is "male", for Age this is "<79years", for Education level this is "higher education", for Financial status this is "easy", for Perceived health status this is "no chronic illness/disability", for Care dependency this is "no care dependency", for Social support this is "high $\geq 3$ ", for Sexual Orientation this is "heterosexual", for Resilience this is "high $\geq 3.40$ (mean)".

Anxiety was associated with lifetime exposure to SV, a higher education, financial difficulties, a chronic illness or disability limiting daily activities, being care dependent, fewer than three confidants, and low resilience. Exposure to SV in the past 12 months, sex at birth, age, and sexual orientation were not associated with anxiety in our sample.

Financial difficulties were associated with PTSD. Lifetime SV was only associated with PTSD in respondents with a chronic illness/disability or no/lower education. Exposure to SV in the past 12 months, sex at birth, age, sexual orientation, care dependency, social support, and resilience were not associated with PTSD in our sample.

The direction of the association between lifetime SV and depression, and anxiety are shown in the first two graphs of Figure 1. The last two graphs show the interaction effect between lifetime SV and education level, and between lifetime SV and perceived health status in the case of PTSD.

**Figure 1.**
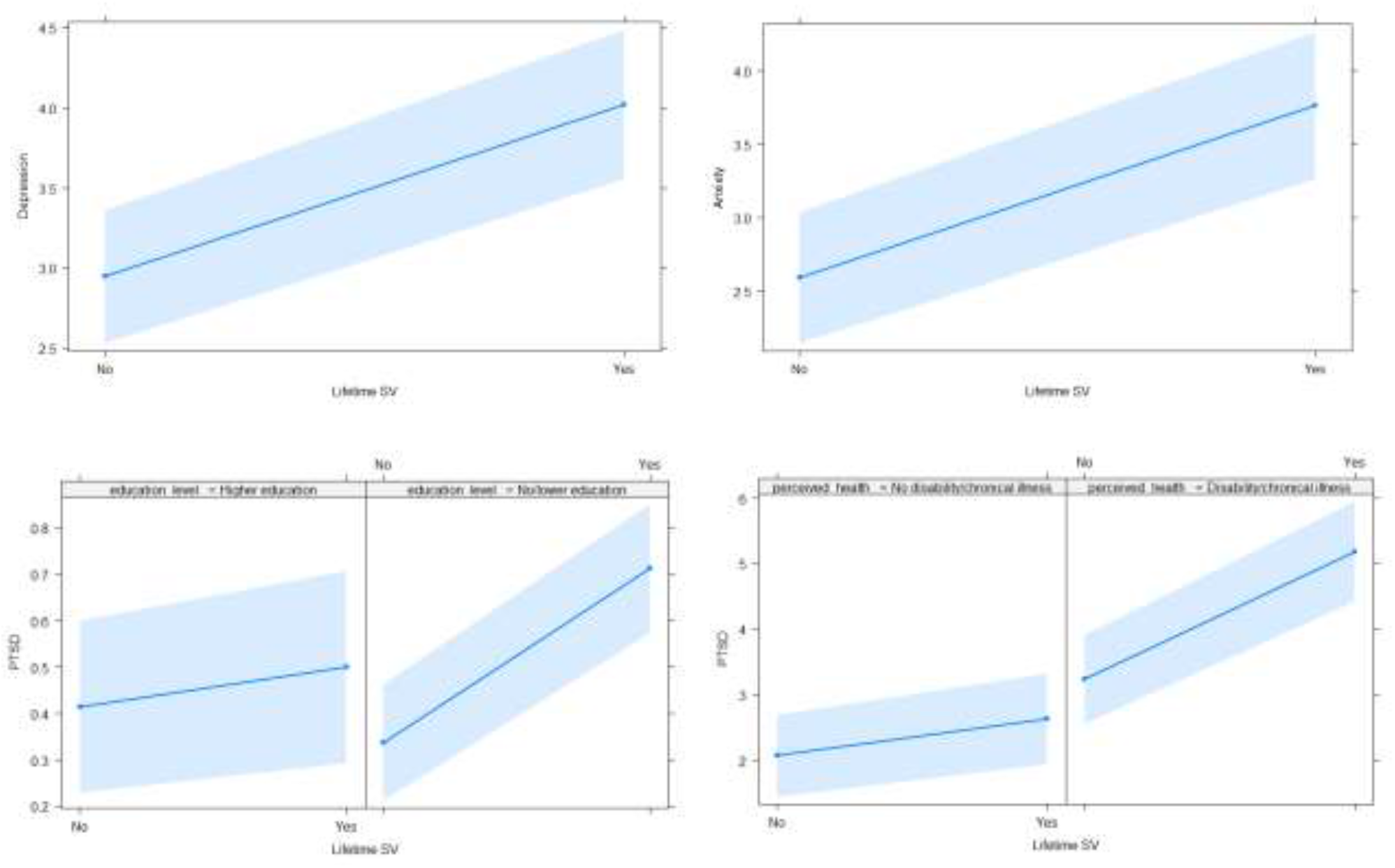
Association between lifetime sexual victimisation with depression, anxiety and PTSD.

## 4. Discussion

In this paper we present the results of a study on the mental health impact of sexual victimisation in a representative sample of 513 older adults living in Belgium. We established the prevalence of depression, anxiety, PTSD, suicide attempts and self-harm in the Belgian populations of 70 years and older. Moreover, we studied the association between lifetime sexual victimisation and depression, anxiety, PTSD, suicide attempts, and self-harm in later life. Furthermore, we examined the moderating effects of sexual victimisation in the past 12-months, sociodemographic characteristics, health status, social support, and resilience in the relationship between lifetime sexual victimisation and depression, anxiety, and PTSD.

### 4.1 Mental health problems are common in older adults in Belgium

Our results confirm that mental health problems are common in older adults (WHO, 2017b). Almost 20% of participants reported mild depressive symptoms, 5.1% reported moderate depressive symptoms, and 2.5% suffered from severe depressive symptoms. Over 17% reported mild, 4.5% moderate, and 3.5% severe anxiety symptoms, and 5.7% suffered from PTSD. Previous studies found similar numbers regarding depression and anxiety (Meeks et al., 2011; Wolitzky-Taylor et al., 2010). However, the PTSD prevalence in our study is almost twice as high compared to previous studies (Böttche et al., 2012). While previous studies defined PTSD according to the criteria of the Diagnostical and Statistical Manual IV (DSM-IV), we used the DSM-5 PTSD criteria, which could influence prevalence rates. Although studies in younger populations showed no difference in prevalence rates when using the DSM-IV versus the DSM-5 criteria (Calhoun et al., 2012; Kilpatrick et al., 2013), this has not yet been researched in older adults. Therefore, more research in older adults comparing different measurement instruments is needed. Regarding suicide attempts and self-harm we found that 1.6% of our study population reported a suicide attempt, and 1.4% self-harm in the past 12-months. Similar rates were found by the Flemish Centre for Suicide Prevention (VLESP) in the same period (Zelfmoord 1813, 2018) Although our results and previous research shows low numbers for suicide attempts and self-harm, it is known that older adults complete suicide proportionally more than any other age group (Rodda et al., 2011; Zelfmoord 1813, 2018). However, information on SV and mental health of older adults who completed suicide were not available in our study.

In addition to the high prevalence of mental health disorders, our study confirms previous findings indicating that financial difficulties, suffering from a chronic illness or disability, being care dependent, and having low social support are associated with adverse mental health outcomes in later life (Cole & Dendukuri, 2003; Rodda et al., 2011; Wolitzky-Taylor et al., 2010). Therefore, we endorse previous calls to invest in psychosocial interventions that combat isolation in older adults, including befriending programmes, and peer support schemes (Wilkinson, Ruane, & Tempest, 2018). Although previous studies link a low education level to adverse mental health outcomes in later life (Cole & Dendukuri, 2003; Rodda et al., 2011; Wolitzky-Taylor et al., 2010), our study shows mixed results. In our sample, older adults with a low education level showed fewer anxiety symptoms compared to highly educated older adults. For depression and PTSD we found no link between education level and symptom severity.

Concerning resilience, our results show that resilient older adults report fewer anxiety symptoms. Whereas previous studies indicated a link between resilience and depressive symptoms in older adults (Hardy, Concato, & Jill, 2004), we found no association in our sample. Since the definition and measurement of resilience continues to be the subject of debate (Leys et al., 2020), and new scales for resilience in older adults were developed and tested very recently (Akatsuka & Tadaka, 2021), the different results could possibly be explained by different measurement instruments. More research comparing the reliability of different measurement instruments for resilience in older adults is needed to draw conclusions concerning its moderating effect on mental health outcomes in later life.

### 4.2. Lifetime sexual victimisation is linked with depression, anxiety, and PTSD in older adults

The second objective of this paper was to check whether lifetime sexual victimisation was associated with depression, anxiety, PTSD, suicide attempts, and self-harm in late life. Both the bivariate and the regression analysis showed an association between lifetime sexual victimisation and current depression, and anxiety in old age. For PTSD we found an association in respondents with a chronic illness/disability or no/lower education. Lifetime sexual victimisation and suicide attempts or self-harm in the past 12-months were not significantly associated. This could be due to the low prevalence rates for suicide-attempts and self-harm limiting the power of our analysis. As discussed earlier, studies show low numbers of suicide attempts in older adults (Zelfmoord 1813, 2018). Since sexual victimisation is a known risk factor for suicide attempts and older adults are more likely to die after a suicide attempt (Rodda et al., 2011; Zelfmoord 1813, 2018), we can partly assume that we could not establish a link between sexual victimisation and suicide attempts in our sample, since possible victims might have died by suicide.

### 4.3 Past 12-months sexual violence, social support, and resilience do not moderate the relation between lifetime sexual victimisation and depression, anxiety, and PTSD

Thirdly, this paper examined the moderating effect of past 12-months SV, sociodemographic characteristics, health status, social support, and resilience in the relation between lifetime sexual victimisation and depression, anxiety, and PTSD in older adults.

In our sample past 12-months SV did not moderate the relation between lifetime sexual victimisation and any of the mental health outcomes, suggesting that exposure to SV earlier in life had such an important impact on the victims’ mental health, that the impact of a recent SV event made no difference. Regarding sociodemographic characteristics we found a moderating effect of education level on the relation between lifetime sexual victimisation and PTSD. Similarly, health status moderates the relationship between lifetime sexual victimisation and PTSD.

Although previous research found that social support in the acute phase after exposure to SV is crucial to decrease adverse mental health outcomes (Declercq & Palmans, 2006; Declercq et al., 2007), our study indicated that social support does not influence long-term mental health outcomes after SV. We assume this could be explained by SV disclosure. Research in younger populations showed that SV victims experience many barriers to disclosure (Dartnall & Jewkes, 2013), which was recently confirmed in a sample of older adults with mental health problems (Nobels et al., 2021b). We hypothesize that social support is only beneficial when the confidants are aware of the victim’s SV history. More research on interaction between social support, SV disclosure and mental health outcomes in older adults is needed to confirm this hypothesis.

Furthermore, resilience did not moderate the effect between lifetime sexual victimisation and depression, anxiety, and PTSD in our sample. Previous studies on the effect of resilience showed mixed results. Some found that resilient subjects may be partly protected against the long-term mental health effect of SV (Schulz et al., 2014), but others could not establish high resilience as a protective factor against adverse mental health outcomes after SV (Fedina et al., 2021). More research on the association between sexual victimisation and resilience on mental health outcomes in older adults is warranted to clarify this.

### 4.4 Limitations

An important limitation of our study was its cross-sectional design, which makes it impossible to establish a causal relation between sexual victimisation and mental health outcomes in later life. Although a longitudinal study described an association between childhood sexual abuse and internalising disorders in old age (Rapsey et al., 2019), another study has shown that people with severe mental illness (SMI) experience substantially more SV during their lifetime compared to those who do not suffer from SMI (Khalifeh et al., 2015). Another limitation was that sexual victimisation was reported retrospectively by the participants. Although there is evidence that the magnitude of associations with mental disorders does not differ according to whether sexual victimisation was reported prospectively or retrospectively (Scott et al., 2012), evidence from a prospective study would further strengthen our conclusions.

### 4.5 Recommendations for future research and clinical practice

Despite its limitations, this study improves the current understanding of the mental health impact of SV in older adults. Being the first to apply a life course perspective to the analysis, including sexual victimisation during childhood, adulthood and old age, and using a broad definition of SV, this study offers a unique understanding of the mental health impact of SV in late life. To strengthen our conclusions, studies using the same approach and SV definition in older adults from different countries and different cultural backgrounds are warranted. Moreover, in order to provide tailored care for older victims of SV, mental health care should be better aligned with the needs of older victims of SV. Therefore, incorporating a routine inquiry about sexual victimisation, as proposed by the WPA (Steward& Chandra, 2017), should become the gold standard of care for older adults who present with mental health problems. However, since health care workers rarely feel comfortable discussing SV with older adults (Goldblatt et al., 2020), capacity building for healthcare workers and the development of clinical guidelines and care procedures seem of the utmost importance.

## 5. Conclusion

Lifetime sexual victimisation is linked to depression and anxiety in older adults in Belgium, and is associated with PTSD in older adults with a chronic illness/disability or no/lower education. Our findings confirm the long-lasting mental health impact of sexual victimisation and show the need for tailored mental health care for older victims of sexual violence. Therefore, more research on the specific needs of older victims of SV regarding mental health care is needed. In addition, professionals working with older adults urgently need capacity building regarding SV and its mental health impact. Furthermore, the development of clinical guidelines and care procedures seems particularly important.

## Data Availability

By request from the corresponding author.

## Acknowledgements

Ines Keygnaert and Christophe Vandeviver contributed equally to this work and are therefore to be regarded as joint last authors of this article.

The authors want to thank Adina Cismaru-Inescu, Lotte De Schrijver, Joke Depraetere, and Laurent Nisen for their input during the questionnaire development. Also, we want to thank Adina Cismaru- Inescu, Bastien Hahaut and Laurent Nisen for their support during the data-collection process. Many thanks to our interviewers for their time and dedication to our study, Elizaveta Fomenko for her statistical advice and Dr. Howard Ryland for his help with the language editing. Finally, we want to thank all older adults who participated in our study for sharing their stories with us.

## 7. Appendix

### Appendix 1. Detailed outcome measurements sexual victimisation

#### Hands-off sexual victimisation (no physical contact)

- *Sexual staring:* Someone stared at me in a sexual way or looked at my intimate body parts (e.g., breasts, vagina, penis, anus) when I didn’t want it to happen.
- *Sexual innuendo:* Someone made teasing comments of a sexual nature about my body or appearance even though I didn’t want it to happen.
- *Showing sexual images:* Someone showed me sexual or obscene materials such as pictures, videos, directly or over the internet (including email, social networks and chat platforms) even though I didn’t want to look at them. This does not include mass mailings or spam.
- *Sexual calls or texts:* Someone made unwelcome sexual or obscene phone calls or texts to me.
- *Voyeurism:* I caught someone watching me, taking photos or filming me when I didn’t want it to happen while I was undressing, nude or having sex.
- *Distribution of sexual images:* Someone distributed naked pictures or videos of me directly or over the internet (including email, social networks and chat platforms) when I didn’t want it to happen.
- *Exhibitionism:* Someone showed their intimate body parts (e.g., breasts, vagina, penis, anus) to me in a sexual way and/or masturbated in front of me when I didn’t want to see it.
- *Forcing to show intimate body parts:* Someone made me show my intimate body parts (e.g., breasts, vagina, penis, anus) online or face-to-face when I didn’t want to do it.

#### Hands-on sexual victimisation

##### Sexual abuse (physical contact but no penetration)

- *Kissing:* Someone kissed me against my will.
- *Touching in care:* Someone touched my intimate body parts (e.g., breasts, vagina, penis, anus) during care against my will.
- *Fondling/rubbing:* Someone fondled or rubbed up against my intimate body parts (e.g., breasts, vagina, penis, anus) against my will.
- *Forced undressing:* Someone removed (some of) my clothes against my will.

#### Rape and attempted rape (physical contact with attempted or completed penetration)

- *Oral penetration:* Someone had oral sex with me or made me give oral sex against my will.
- *Attempt of oral penetration:* Someone tried, but did not succeed, to have oral sex with me or tried to make me give oral sex against my will.
- *Vaginal or anal penetration:* Someone put their penis, finger(s) or object(s) into my vagina or anus against my will.
- *Attempt of vaginal or anal penetration:* Someone tried, but did not succeed to put their penis, finger(s) or object(s) into my vagina or anus against my will.
- *Forcing to penetrate:* Someone made me put my penis, finger(s) or object(s) into their (or someone’s) vagina or anus against my will.

